# The Neuroendocrine Profile During the Trier Social Stress Test in College Freshmen Offers Insights into the Emergence of Anxiety and Depression Symptoms

**DOI:** 10.64898/2026.01.27.26344962

**Authors:** Huzefa Khalil, Cortney A Turner, Virginia Murphy-Weinberg, Linda Gates, Fei Li, Alexandra Onica, Keiko Arakawa, Lauren Weinberg, Clare Stack, Juan F Lopez, Stanley J Watson, Huda Akil

**Affiliations:** Michigan Neuroscience Institute, University of Michigan, Ann Arbor, MI, USA; Department of Pharmacology, University of Michigan, Ann Arbor, MI, USA; Department of Psychiatry, University of Michigan, Ann Arbor, MI, USA

## Abstract

**Background:** The Michigan Freshman Study on Stress and Resilience aims to identify factors that predict the emergence of depression and/or anxiety symptoms in college freshmen. We previously showed that a combination of psychiatric instruments (Affect Score) strongly predicts who will develop such symptoms during the freshman year. Here, we ask: a) Can we replicate the predictive power of the Affect Score in an independent cohort? and b) Can the neuroendocrine profile during the Trier Social Stress Test (TSST) serve as an additional predictor?

**Methods:** A new cohort of subjects (N= 357) was used for Affect Score replication. The TSST study involved 337 subjects (Females 184, Males 153). Self-report questionnaires at the start of the year were used to derive the Affect Score. GAD-7 and PHQ-9 were used to monitor anxiety and depression, respectively. TSST measures involved plasma ACTH and Cortisol and heart rate monitoring.

**Results:** The Affect Score proved to be a highly replicable predictor of future depression and anxiety. In the TSST, subjects not currently depressed but who developed depression at another timepoint during the year showed a higher and delayed peak of the CORT response. Female subjects not currently anxious but who developed anxiety at another timepoint had an elevated CORT response throughout the TSST. This hyperresponsiveness was not correlated with Affect Score and was an independent predictor of anxiety.

**Present address:** Michigan Neuroscience Institute, University of Michigan, A. Alfred Taubman Biomedical Science Research Building, Rm 2009, Ann Arbor, MI, 48109-9901, USA

**Author Contributions:** HK performed research, analyzed data, wrote the paper; CAT designed research, performed research, wrote the paper; VM-W designed research, performed research; LG, FL, AO, KA and LW performed research; CS coded and analyzed data; JFL designed research; SJW Jr designed research; HA designed research, wrote the paper.

**Funding:** This work was supported by the Office of Naval Research (ONR) Grant N00014-09-1-0598, N00014-12-1-0366 and N00014-19-1-2149, the Pritzker Neuropsychiatric Disorders Research Consortium Fund, LLC and the Hope for Depression Research Foundation. This project was also supported by Grant Number P30DK020572 (MDRC) from the National Institute of Diabetes and Digestive and Kidney Diseases.

**Competing interests:** The authors declare no competing interests.

**Conclusions:** The Affect Score is a powerful predictor of depression and anxiety in college freshmen. The combination of Affect Score and TSST is strongly predictive of anxiety in females.

## Introduction

The Michigan Freshmen Study on Stress and Resilience aims to predict the emergence of depression and anxiety symptoms in a young and healthy cohort of college freshmen by using a combination of biological and psychological variables at the start of the freshman year and following the students through the academic year and into the fall of their sophomore year. The goal is to identify the risk variables whereby the stress of the college experience triggers significant symptoms of depression and anxiety.

We previously reported that the freshman year was a reliable trigger of increased depression and anxiety across multiple cohorts. During the Covid-19 pandemic, baseline rates of depression and anxiety rose steadily, and were further exacerbated during the freshman year with over 33% of the subjects meeting criteria for depression (>40% in females) (Turner et al., 2023). In that study, we also used machine learning to derive a composite index called the “Affect Score”, which included baseline psychological state and trait measures along with family history. This index proved to be a strong predictor of depression symptoms experienced during the freshman year (Turner et al., 2023). Indeed, the Affect Score predicted depression far better than the polygenic risk score for depression (MDD-PRS) and did so before and during the Covid-19 pandemic. But it is critical to ascertain whether this index is reliable in a cohort that is independent of the one from which it was derived. Thus, one goal was to test the predictive power of the Affect Score in a new cohort of subjects.

A second question was whether any biological indices, beyond genetics, were predictive of the emergence of depression and/or anxiety in this population. Thus, we focused on the neuroendocrine response to a social stressor, the Trier Social Stress Test (TSST), to explore its usefulness in predicting susceptibility or resilience to stress. Our working hypothesis is that a healthy endocrine stress response is adaptive and is characterized by a rapid rise of adrenocorticotropic hormone (ACTH) in the blood, followed by a rise in cortisol (CORT) which triggers fast feedback and results in the rapid termination of the endocrine response (McEwen and Akil, 2020). By contrast, mood and anxiety disorders are often associated with either an exaggerated or blunted stress response (Kendler et al., 1993; Kessler, 1997; McEwen, 2000; McEwen and Akil, 2020; Akil, 2005), and dysregulation of negative feedback, which typically leads to adverse outcomes (McEwen, 2000; Sapolsky, 1996). We refer to the ability of the individual to cope with ongoing stress and exhibit a strong but time-limited physiological stress response as “Stress Fitness”. *Here we ask whether the endocrine profile to a social stressor is a useful descriptor of “stress fitness”, and whether its dysregulation is predictive of future emergence of mood and anxiety symptoms in our population*.

The TSST is an acute laboratory challenge comprised of a speech task and an arithmetic task that the subject performs in front of a panel of three judges. A number of studies have provided evidence of a dysregulated neuroendocrine response during the TSST in depressed patients (Young, Lopez, et al., 2000; Gotthardt et al., 1995; Trestman et al., 1991; Man et al., 2022) or during a similar stress paradigm that combines elements of the TSST and the Cold Pressor Test, known as the Maas- tricht Acute Stress Test (Kuhn et al., 2025). Notably, patients with comorbid depression and anxiety show a significantly higher ACTH response relative to controls and those with isolated depression or anxiety, indicating an association between comorbidity and increased HPA axis activation under stress (Young, Abelson, and Cameron, 2004). Another important biological indicator is heart rate and heart rate variability, which have also been shown to be dysregulated in patients with current depression, anxiety or their combination (Schiweck et al., 2019; Seipäjärvi et al., 2022; Carney and Freedland, 2009). It should be noted that sex differences have been observed during the TSST both in endocrine response (Kirschbaum, Kudielka, et al., 1999) and heart rate variability (Koenig and Thayer, 2016; Hamidovic et al., 2020).

While the evidence of a dysregulated TSST response is strong in clinical populations, the TSST profile has not been used as a predictive tool in individuals not currently suffering from depression or anxiety. To explore this potential, we measured heart rate, blood CORT and ACTH levels before, during and after the social task. We asked

a. What is the stress profile in subjects who are currently exhibiting symptoms of depression and anxiety?
b. In subjects *without current or depression or anxiety*, do any components of the stress response *predict* the incidence of depression or anxiety at other time points?
c. Are the stress predictive parameters associated with the Affect Score or independent of it?
d. Are there sex differences in these relationships?

## Methods & Materials

### Subjects

During academic years from 2015–19, and from 2023–24 we recruited incoming freshmen at the University of Michigan for the TSST (N=337). Of the 184 females and 154 males who enrolled, 317 subjects completed follow-up questionnaires (176 females, including 27 on birth control, and 141 males). The demographics of the subjects are shown in Supplemental Table 1. We recruited according to the methods in Turner et al., 2023. Subjects were recruited via email, Facebook and posted flyers. All subjects provided written informed consent. The protocol was approved under HUM00028766 and HUM00211856 by the University of Michigan Medical School Institutional Review Board. For the first four years, subjects who took birth control (BC) were excluded, but that criterion was relaxed in later years to increase enrollment. Saliva samples were also collected for various other measures during the year. Hair samples were also collected for some of the years for other analyses.

**Table 1.**
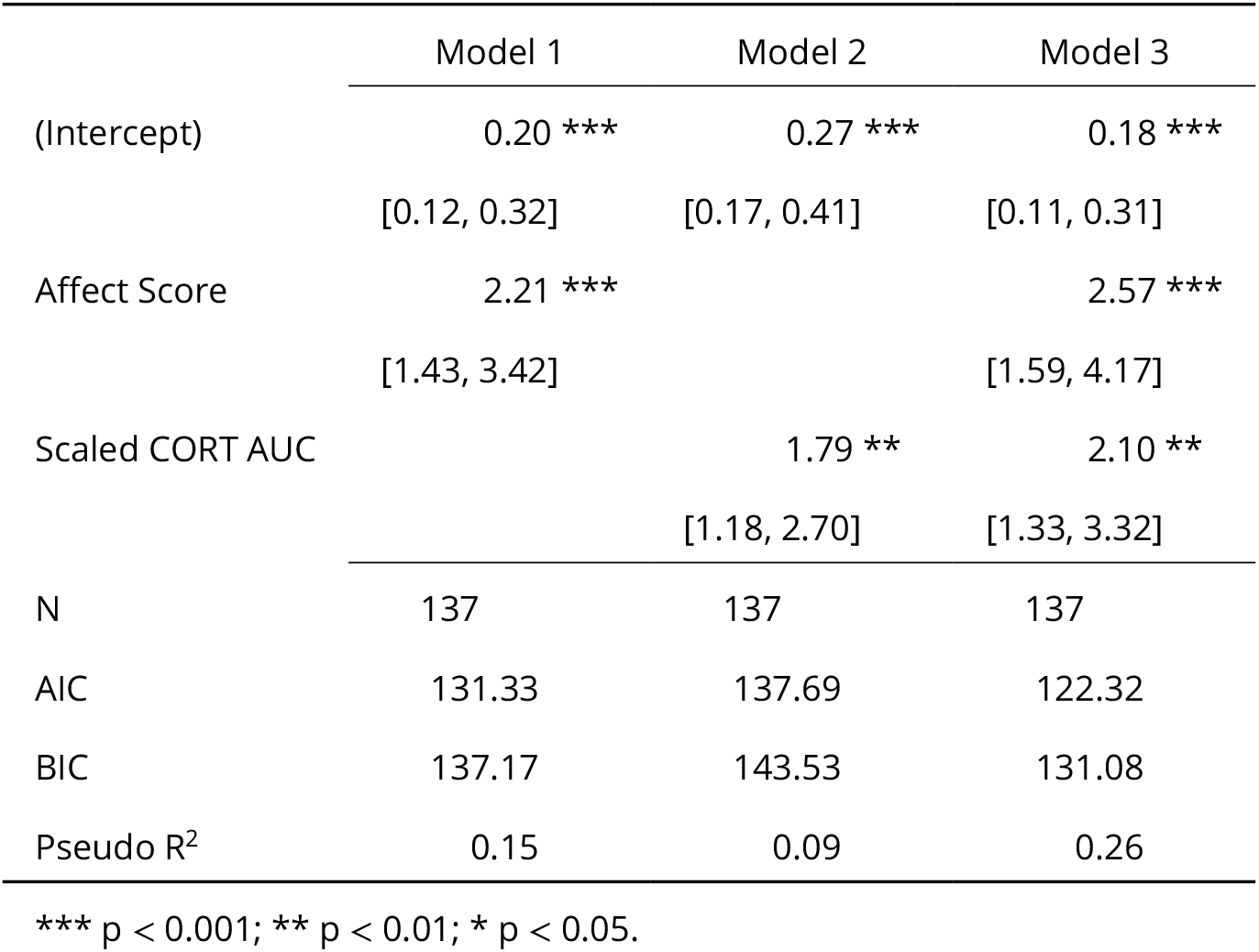
Full TSST CORT response and Affect Score. Prediction of follow-up anxiety for female subjects not on birth control. The dependent variable is binary yes / no follow-up anxiety.

While we recruited subjects from 2020–2023, we could not conduct the TSST because of inperson research restrictions during the COVID-19 pandemic. Additionally, there were subjects in the 2023 and 2024 cohorts who did not undergo the TSST. Data from these additional subjects (N = 289, 182 female) is shown in the context of the replication of the Affect Score.

### Self-report Questionnaires

At baseline, we administered a variety of questionnaires at the start of the freshman year (June-September of each year). The methods are the same as from Turner et al., 2023. This battery included the General Anxiety Disorder-7 (GAD-7) for anxiety symptoms and the Patient Health Questionnaire-9 (PHQ-9) for depression symptoms. The subjects were given the following 12 additional questionnaires (14 measures): the NEO Personality Inventory-Revised (NEO PI-R), the Risky Family Questionnaire (RFQ), the Childhood Trauma Questionnaire (CTQ), the Spielberger State-Trait Anxiety Inventory (SB State, SB Trait), the Positive and Negative Suicide Ideation Inventory (PANSI+, PANSI-), the Morningness-Eveningness Questionnaire (MEQ), the Perceived Stress Scale- 10 (PSS), the Multidimensional Scale of Perceived Social Support (MSPSS), the Connor-Davidson Resilience scale (CD-RISC), the 5-item Dispositional Positive Emotions subscale (Compassion), the Pearlin Mastery scale (Mastery), and the 3-item Revised UCLA Loneliness scale (Loneliness). Both Qualtrics and RedCap were used to collect information over the years, and some scales could only be administered on paper.

At follow-up, the same anxiety and depression questionnaires were administered via Qualtrics or RedCap at four follow-up time points (3-months, 6-months, 9-months, and 12-months) after the start of the freshmen year. Other questionnaires were also administered either quarterly or triannually. The highest follow-up GAD-7 or PHQ-9 value was used to determine the presence of anxiety of depression symptoms during the school year, respectively. For classification purposes, we used a cutoff value of 10, as this cut-off has been shown to have high sensitivity and specificity for both tests. Any subject missing more than two (50%) of the follow-up time points was excluded from any analysis related to anxiety or depression during the freshmen year.

### Trier Social Stress Test (TSST)

The TSST was performed according to previous methods (Kirschbaum, Pirke, and Hellhammer, 1993; Mayer et al., 2020; Young, Abelson, and Cameron, 2004). The test was conducted on students during the fall term for all academic years, except 2019, 2023 and 2024 when the TSST went through April. Briefly, subjects were inserted with an IV catheter between 2-3pm. After 60 minutes, the subjects were brought to a different room and asked to prepare for a public speaking task (time 0) in front of a panel of 3 individuals (with male and female panelists). At 5 minutes, the subjects began the public speaking task. At 10 minutes, the subjects were asked to count backwards from 1022 subtracting 13. The subject then left the room, and the IV remained in until 65 minutes after the start of the TSST (time 0). The blood was collected in EDTA BD vacutainer tubes that were kept on ice and spun within 15 minutes of collection at 3300 rpm and 4°C for 10 minutes. The plasma was then aliquoted into 1 mL aliquots and stored at -80°C until processing.

### Hormone Analyses

Adrenocorticotropic hormone (ACTH) and Cortisol (CORT) were analyzed by the Michigan Diabetes Research Center Chemical Laboratory at the University of Michigan. The assay was a chemiluminescent assay using the Siemens Immulite 1000 automated analyzer. Cortisol levels were in µg/dL, and ACTH levels were in pg/mL. For 2024–25, the research core purchased a Siemens Immulite 2000 XPi with a lower limit of detection for ACTH, and a correction was needed to make the results comparable to the previous years. The ACTH values were corrected by calculating a correction factor for each time point, averaging these factors and then applying this correction factor to the data from 2024-25.

### Fitbit

All subjects wore Fitbits (Charge HR, Charge 2, Charge 3, Inspire 2 or Inspire 3). Data were collected by either Fitabase or CareEvolution. Heart rate (HR) was collected before, during and after the TSST session. The heart rate data is shown as beats per minute (BPM).

### Statistical Analyses

All statistical analyses were performed in R v4.5.1 (R Core Team, 2025). A Mann-Whitney U test was done for unpaired tests, and a Wilcoxon signed rank test was done for paired tests to test for differences between groups. The Area Under the Curve (AUC) was calculated for the TSST in the following time intervals in minutes: Pre-TSST (-60 to 0), TSST (0 to +15), Post-TSST (0 to +65) and the Full TSST (Pre-TSST + TSST + Post-TSST). The average Pre-TSST CORT response, the Full TSST CORT AUC response and the rise in CORT during the TSST (0 to +15) were calculated for indepth analyses. Pearson’s correlation was used for all correlations shown. Logistic regression was used to predict follow-up anxiety. A repeated measures ANOVA, calculated using a random-effects model, was used when appropriate. The p-values for odds ratios are from Fisher’s exact test. The Affect Score is the z-score of the first dimension of the PCA of 10 baseline surveys selected using machine learning (Turner et al., 2023).

## Results

### Depression and anxiety during the freshmen year

The subject demographics are shown in Supplemental Table 1. At baseline, 4.4% and 6.2% of subjects had depression and anxiety symptoms, respectively. During the freshmen year, 23.7% and 21.7% of subjects developed depression and anxiety symptoms, respectively. Even though this data spans many years before, during and after the pandemic, including more recent years, the results were comparable to our previously published results when sampled quarterly (Turner et al., 2023).

### The Affect Score replicated its power in predicting follow-up depression and anxiety scores

A separate cohort of subjects (N=357), independent from the cohort used to derive the Affect Score, was used to determine whether the Affect Score continued to be predictive of follow-up depression and anxiety. Here again, the Affect Score proved highly significantly correlated (*r*=0.69, *p*<0.001) with follow-up depression symptoms (Fig. 1A). Moreover, the odds ratio of developing depression with an Affect Score greater than 0 was 14.25 (*p*<0.001). This replicates our findings that the Affect Score is a significant predictor for developing depression during the freshmen year. Similarly, the

**Figure 1.**
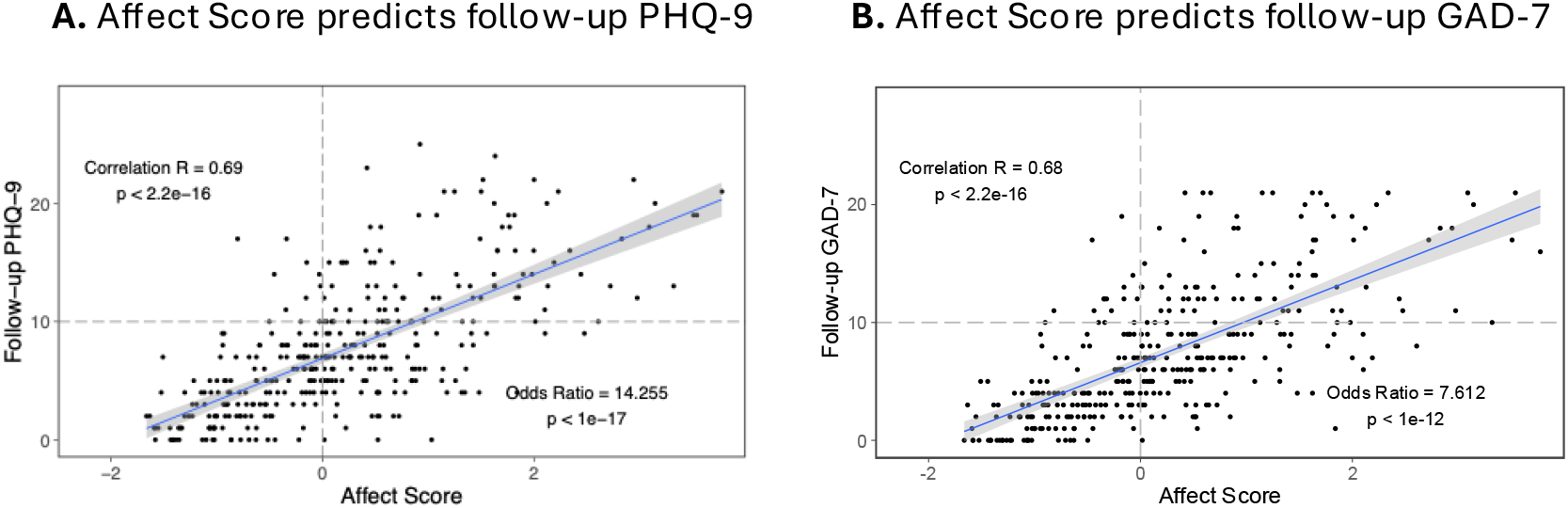
Replicating the ability of the Affect Score to predict follow-up depression (A) and anxiety (B) scores. The predictive power of our previously derived Affect Score was replicated in a new, independent cohort of freshmen for the four academic years of 2021-2024 (N=357).

Affect Score has a strong predictive power for anxiety as seen in Fig 1B, with a correlation of 0.68 (*p*<0.001) and an odds ratio of 7.6 (*p*<0.001).

### The TSST response is altered in females on birth control

We first looked at the effect of sex and birth control on the TSST response. Figure 2A shows the response of all the subjects to the TSST for both ACTH and CORT. There was a clear increase in ACTH upon IV insertion, as well as at the start of the TSST. This was followed by an increase in CORT which had a later peak with a longer turnoff period to for returning to baseline. Overall, the stress response in this population reflected a classic, healthy response to an acute challenge.

**Figure 2.**
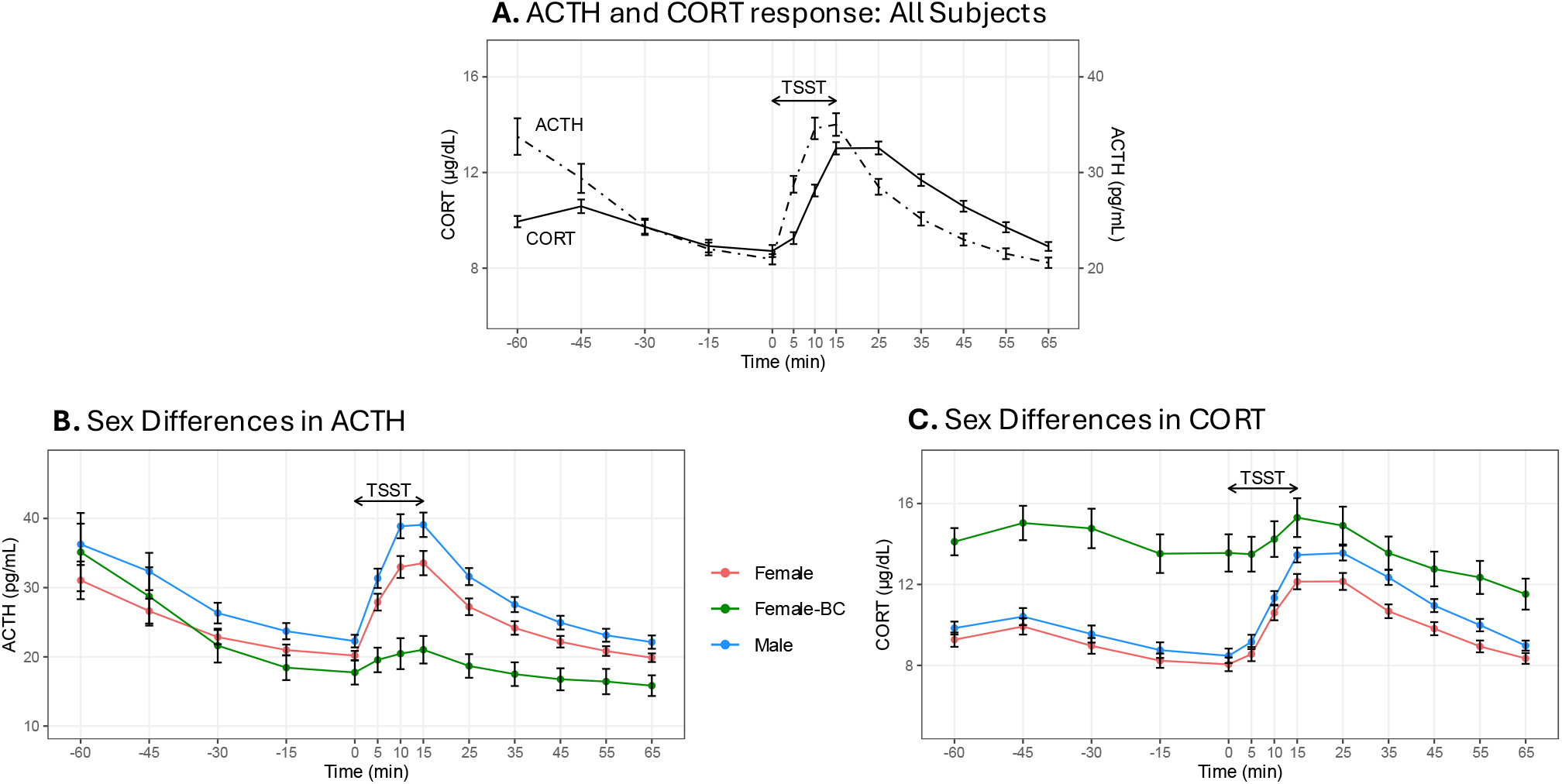
The physiological stress response to the TSST: Effect of sex. A) The CORT and ACTH response to the TSST. Time 0 is the start of the TSST and Time 15 is the end of the TSST as shown by the arrow The IV is inserted 60 minutes before the TSST and the last blood sample is collected 65 minutes after the start of the TSST. B) The ACTH response was blunted in females taking birth control when compared to females not on birth control (p < 0.001) and higher in males as compared to females not on birth control (p = 0.003). C) The CORT response was higher in females taking birth control than in females not on birth control (p < 0.001), and the females taking birth control had a decreased rate of rise during the TSST. The TSST CORT response was higher in males when compared to females not on birth control (p = 0.02). Female N = 155, Female on birth control N = 29, Male N = 153. p-values from a Wilcoxon rank-sum test of group differences in the Area Under the Curve (AUC).

In Figure 2B, the ACTH response is broken out by sex, with females subjects on BC plotted separately. Similarly, Figure 2 C shows the corresponding CORT response separated by males, females and females on BC. There is a clear and significant difference in both the ACTH and CORT curves for females on BC as they show a blunted ACTH response and a high CORT response throughout. Given these large differences due to BC, female subjects on BC were excluded from further analyses. In addition, males had higher ACTH and CORT levels than females in general, though they both followed a similar pattern.

### Depression symptoms are associated with a blunted neuroendocrine response during the TSST

We then assessed the TSST response in individuals who exhibited anxiety or depression symptoms at the time of the TSST.

#### Current Depression

In Figure 3A, we see that the ACTH response was blunted in individuals with depression symptoms at the time of the TSST. The CORT response shown in Figure 3B was elevated prior to the TSST but was blunted during the social stressor (i.e. the speaking and math tasks) (p<0.05). Moreover, the rise in CORT levels during the TSST was inversely correlated with PHQ-9 scores (Supplemental Fig 1).

**Figure 3.**
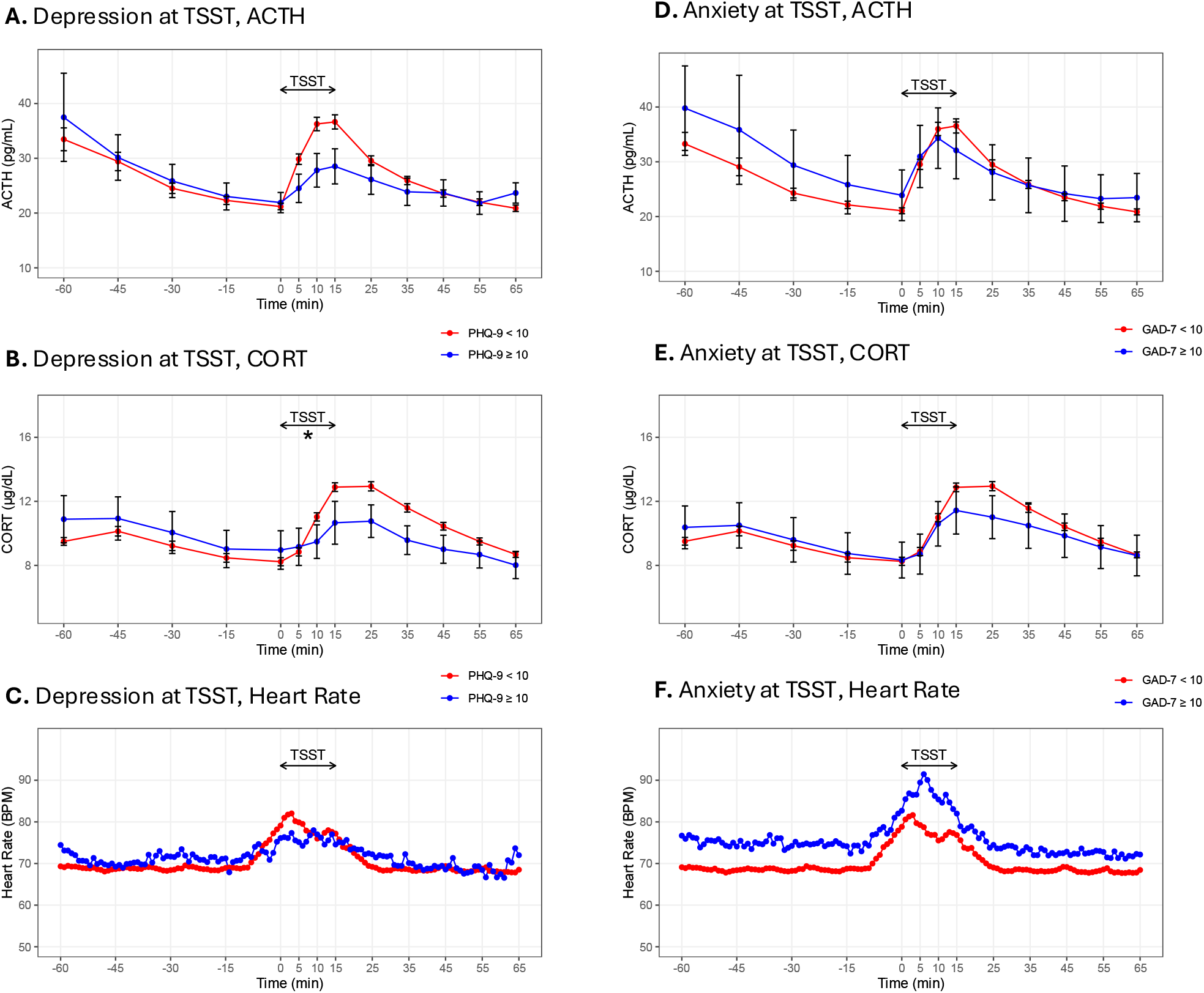
Key variables that modify the physiological stress response to the TSST: The effect of current depression or anxiety. Subjects with depression (PHQ-9 ≥ 10) at the time of the TSST showed a blunted ACTH (A) and CORT (B) response to the TSST whereas there was no difference in the Heart Rate (C). The rise in CORT was significantly lower in depressed subjects (p = 0.024). There was no difference in the ACTH (D) and CORT (E) responses of subjects with or without anxiety (GAD-7 ≥ 10) at the time of the TSST while subjects with anxiety showed a higher heart rate (F). For A and B, No Depression N = 295 (148 Female) and Depression N = 13 (7 Female). For C, No Depression N = 258 (134 Female) and Depression N = 9 (5 Female). For D and E, No Anxiety N = 291 (145 Female) and Anxiety N = 17 (10 Female). For F, No Anxiety N = 254 (131 Female) and Anxiety N = 13 (8 Female).

*Heart rate* was not different in those with and without depression symptoms at the time of the TSST (Fig 3C).

#### Current Anxiety

By contrast, there were no significant differences in the ACTH or CORT response between subjects who did and did not have anxiety at the time of the TSST (Figures 3D and 3E).

*Heart Rate* was consistently higher in subjects with anxiety symptoms (Fig 3F, p = 0.051).

### The CORT response and follow-up depression and anxiety symptoms

To understand if the neuroendocrine response can *predict* anxiety or depression symptoms, we first excluded all subjects with PHQ-9 or GAD-7 scores of 10 or more at the time of the TSST from any further analyses. Figure 4 shows the ACTH and CORT responses for the remaining subjects separated by follow-up depression or anxiety symptoms developed at some point during the freshman year.

**Figure 4.**
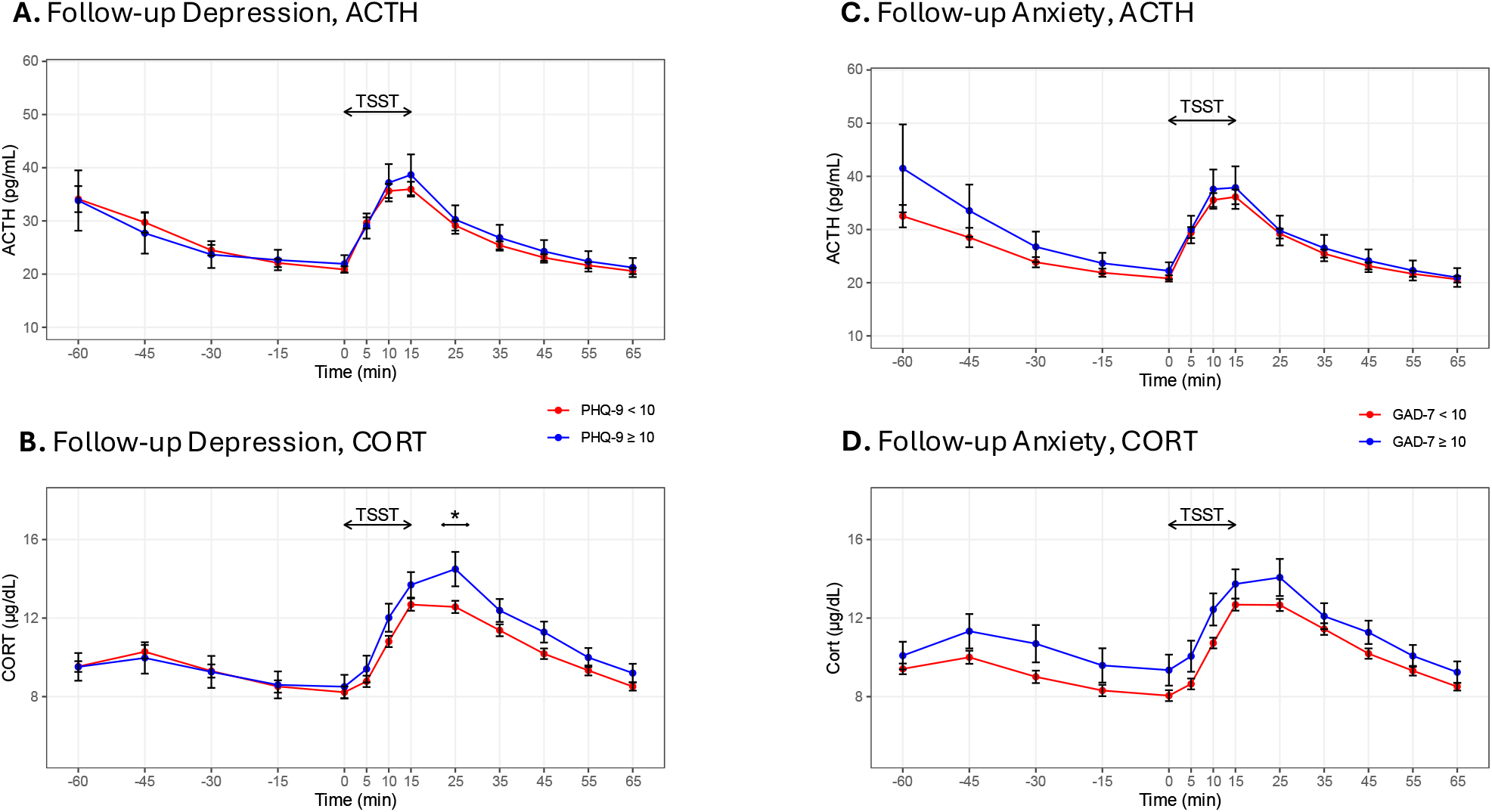
Elevation of the CORT response is associated with the development of follow-up anxiety symptoms in those without current anxiety. The ACTH response (A) showed no differences in subjects who develop depression symptoms at follow-up while the CORT response (B) was slightly elevated post-TSST (p = 0.029) in these subjects. The ACTH response (C) was not different in those who develop anxiety at follow-up while the CORT response (D) was significantly elevated in those who develop anxiety, F(1, 267) = 4.81, p = 0.029. For A and B, No Follow-up Depression N = 222 (110 Female) and Follow-up Depression N = 47 (27 Female). For C, D, No Follow-up Anxiety N = 223 (108 Female) and Follow-up Anxiety N = 46 (29 Female).

#### Follow-Up Depression

Fig 4A shows that there were no between group differences in the ACTH response between subjects who never met criteria for depression and subject who exhibited depression at other timepoints. By contrast, Fig 4B shows that the CORT response does detect differences between these two groups. Thus, subjects who develop depression reached the CORT peak after the end of the TSST indicating that CORT continued to increase even after the social stressor ended. This was not observed in subjects who did not develop depression symptoms and as such, we see a significant difference in CORT levels at 25min (p = 0.029). This pattern of responsiveness is suggestive of some dysregulation in fast feedback resulting in an increase in the duration of the CORT response and slightly elevated CORT during the waning component of the stress response.

There is also evidence of heterogeneity in the endocrine patterns associated with the severity of follow-up depression. Indeed, if we categorize the subjects with follow-up PHQ-9 scores as having no depression (PHQ-9 < 10), moderate depression (PHQ-9 between 10 and 15) and severe depression (PHQ-9 ≥ 15), there were differences in the ACTH and CORT profiles between the three groups (Supplemental Fig 2). In particular, subjects with severe follow-up depression start off with high ACTH and CORT levels and do not exhibit as sharp of a rise or turnoff in response to the social stressor but a flatter, broader and highly dysregulated response. While this did not reach significance due to the small number of subjects with exhibited severe depression at other time points (N=11), it is highly suggestive and merits further investigation.

#### Follow-Up Anxiety

As shown in Fig 4C there was no significant difference in the ACTH response in subjects who develop future anxiety.

Figure 4D shows that subjects who develop anxiety at some point during the freshman year had higher CORT levels throughout. A repeated measures ANOVA found this difference to be significant (Supplemental Table 2). Breaking this up by sex shows that the higher CORT in subjects with followup anxiety is driven by female subjects (Fig 5B), with no significant difference observed in male subjects (Fig 5A).

**Figure 5.**
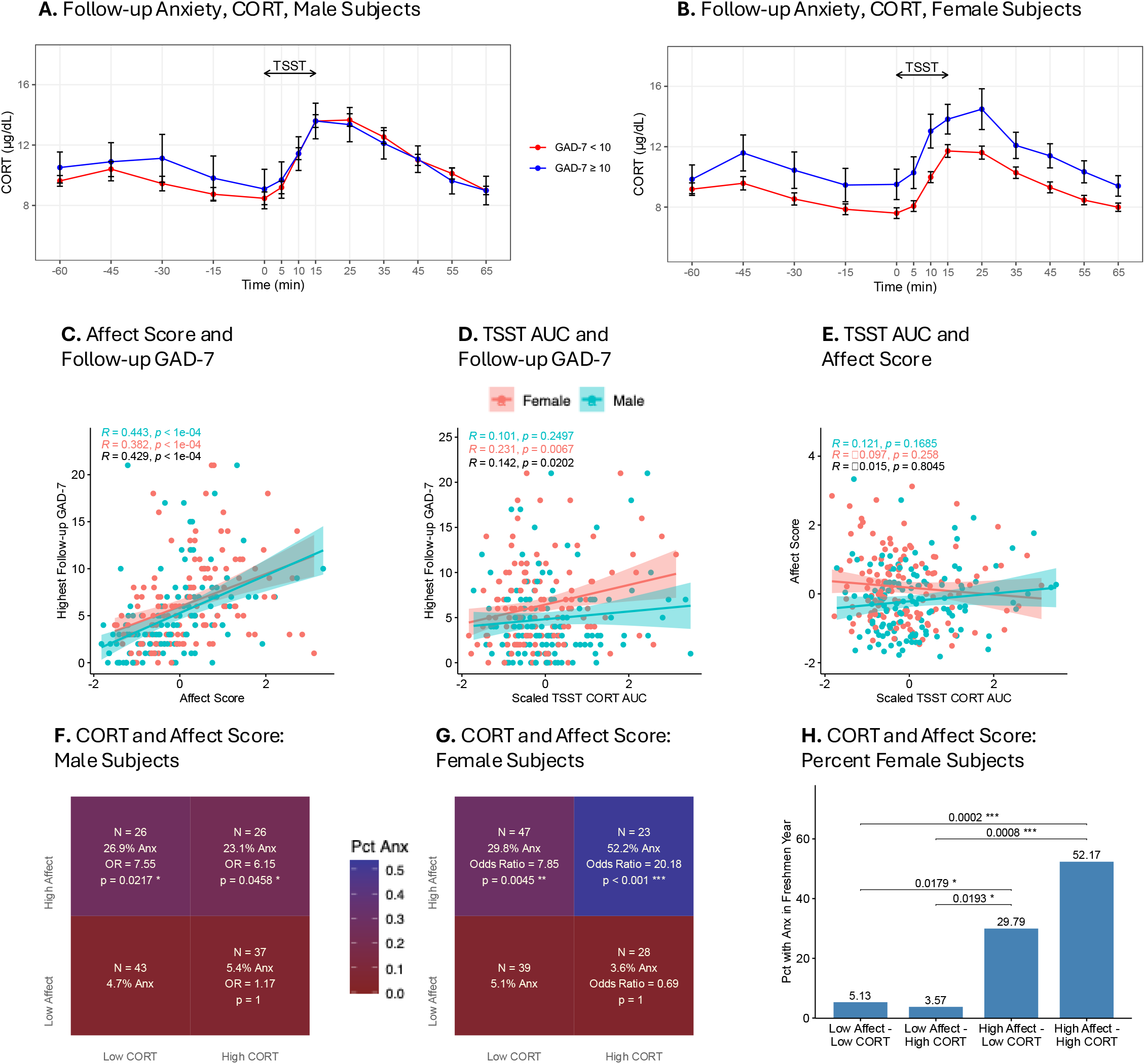
Predicting the emergence of anxiety: Affect Score and CORT response profile in females not on birth control and without current anxiety symptoms. A) CORT response profile for female subjects showed elevated CORT levels for those subjects who met criteria for anxiety at some point during the Freshmen year, F(1, 134.91) = 9.08, p = 0.0031. B) There was no difference for male subjects. C) The Affect Score was positively correlated with follow-up anxiety symptoms in the TSST subjects. D) The scaled CORT response AUC was correlated with follow-up anxiety in female and male subjects. E) The scaled CORT response AUC was not correlated with the Affect Score. F) High CORT was not additive in increasing the risk of anxiety for male subjects but G) was so for female subjects. H) Percent with anxiety in each group with adjusted p-values from Fisher’s Exact Test showing differences between groups. For A, N = 137 (29 with anxiety). For B, N = 132 (17 with anxiety). For C, D and E, N Female = 137, N Male = 132

#### The interaction of Affect Score and CORT levels in predicting anxiety

We have now shown in two independent cohorts that the Affect Score is highly predictive of follow-up anxiety and depression. Since the subjects used in the TSST study are a subset of the subjects from both the original cohort (Turner et al, 2023) and the new cohort, we asked whether the TSST group also shows a similar pattern. Figure 5C confirms the correlation between the highest follow-up GAD-7 score and the Affect Score for the TSST subjects, showing a highly significant correlation for both male and female subjects.

We then asked: Do neuroendocrine measures during the TSST add to the power of the Affect Score in predicting anxiety? In Figure 5D, we calculated the Area Under the Curve (AUC) of the CORT response and found that it was positively correlated with follow-up GAD-7 scores. This correlation was significant for both female and male subjects and indicates that those with higher GAD-7 scores during the freshman year exhibit higher CORT levels during the TSST.

We then asked whether this relationship between the CORT response and anxiety levels was mediated by the Affect Score. Figure 5E shows that there was no correlation between the Affect Score and CORT levels during the TSST, suggesting that CORT levels capture a dimension that is not captured by the Affect Score in predicting anxiety levels.

Given these findings, we classified our subjects into those with high / low Affect Score and high / low CORT levels (levels above and below the mean). In both male and female subjects, a high Affect Score was very predictive of anxiety as the odds ratio rises to around 7. High CORT, by itself, was not predictive of anxiety—i.e., if subjects have a low Affect Score, high CORT did not increase the odds of getting anxious. However, the combination of high Affect and high CORT was highly predictive, and especially notable in female subjects. As shown in Fig 5G, in females with high affect score and high CORT, the odds of developing anxiety are ∼20 fold higher than the odds for females with low Affect Score and low CORT. Indeed, more than half of all female subjects with both high CORT and high Affect developed anxiety symptoms at some point during the freshmen year (Fig 5H). The additive effect of high CORT was not observed for male subjects, as the odds ratio remained the same regardless of CORT levels (Fig 5F). There was a smaller additive effect of high CORT and high Affect in the case of depression in males (Supplemental Fig 3).

Similar conclusions emerge from the results of three binomial regression models for follow-up anxiety for female subjects in Table 1. In Model 1, one standard deviation increase in the Affect Score increased the odds ratio of anxiety by 2.22. In Model 2, a corresponding one standard deviation increase in CORT increased the odds ratio of anxiety by 1.79. Model 3 includes both the Affect Score and CORT, and the model explained approximately 26% of the variance (pseudo R2), and both the Affect Score and CORT levels were highly significant predictors of anxiety.

## Discussion

This study further explored the factors that predict the emergence of depression and/or anxiety in a young cohort of subjects exposed to a naturalistic stressor. We asked whether a previously identified algorithm continued to be a valid predictor. We also asked whether a dysregulated stress response was predictive of susceptibility to depression and/or anxiety.

The main findings can be summarized as follows: A) The Affect Score replicated its ability to predict follow-up depression and follow-up anxiety. B) Female subjects on birth control showed a blunted ACTH response to the stressor while having very high CORT levels throughout. C) Subjects depressed at the time of the TSST had a blunted CORT response. Moreover, the rate of rise in CORT levels due to the stressor was negatively correlated with PHQ-9 scores indicating decreased reactivity of the neuroendocrine stress response. D) Those who met criteria for anxiety had higher heart rate throughout the TSST. E) The CORT response during the TSST proved informative in subjects not currently anxious or depressed but who met criteria for depression or anxiety at some other point during the freshman year: Subjects who became depressed exhibited a later and higher peak in CORT levels relative to those who never became depressed. And a subset of these subjects who met criteria for severe depression had higher basal CORT levels with a lower rise in CORT due to the social stressor. The most consistent finding was in female subjects who were not currently anxious but met criteria for anxiety at another timepoint. They exhibited consistently elevated CORT levels throughout the TSST. CORT levels were not correlated with the Affect Score, indicating that these two variables capture two different dimensions in the prediction of anxiety. The combination of high CORT and high Affect was particularly powerful in predicting anxiety in female subjects. For male subjects, high CORT and high Affect was additive in predicting depression, but the effect was not as strong as that seen in the case of anxiety in females.

In this study, we replicated the finding that the Affect Score is highly predictive of both depression and anxiety levels in an independent cohort. This index consists of selected items from various psychological and psychiatric instruments that capture psychological state and trait measures as well as family history, derived using machine learning in an earlier cohort (Turner et al., 2023). The Affect Score explains approximately 50% of the variance in predicting risk for depression, which is much higher than the risk explained by MDD-PRS (5.8% in people with European ancestry in Adams et al., 2025) and has proven useful regardless of sex or ethnicity.

Overall, this young population exhibited a classic pattern of neuroendocrine stress responsiveness consistent with “stress fitness”- a sharp rise in ACTH followed by a rise in CORT, with high CORT levels leading to a shutdown of ACTH followed by a turn off in CORT, indicating effective negative feedback. There were some sex differences in the response profiles. Males had higher ACTH than females in response to the TSST, as well as a slightly higher CORT response compared to females not on birth control. This was also observed in a meta-analysis of the salivary cortisol response to the TSST and suggests that the females not on birth control may have lower ACTH and CORT levels due to a portion in the follicular phase (Gu et al., 2022). However, females on birth control had a blunted ACTH response and a higher overall CORT response to the TSST. This pattern of females on birth control was similar to other data in plasma (Kirschbaum, Kudielka, et al., 1999). By contrast, researchers have found that the salivary CORT response was blunted on birth control (Bouma et al., 2009; Gervasio et al., 2022). One possible explanation for our finding is that cortisol binding globulin in blood is increased by BC, leading to a constitutively enhanced activation of the CORT response (Kumsta et al., 2007), and suggesting that females on birth control may not mount an ACTH response due to negative feedback induced by high plasma CORT (Gervasio et al., 2022). Depression symptoms at the time of the TSST resulted in a blunted stress response. In general, these individuals did not mount a typical TSST response, as the rise in CORT was lower during the TSST. Moreover, the rate of CORT rise was negatively correlated with PHQ-9 scores, suggesting that higher current depression leads to a blunting in the CORT response. This is at odds with some of the reports in the literature in depression where depressed individuals are hyperresponsive, not hyporesponsive (Carroll et al., 2007). However, these studies were conducted on severely and chronically depressed individuals with melancholic or psychotic depression. In our study, most of the subjects were moderately depressed and exhibited an atypical neuroendocrine profile indicative of a possible lack of stress fitness. By contrast to current depression, current anxiety during the TSST did not alter the neuroendocrine stress response. However, heart rate was increased in those with anxiety symptoms at the time of the TSST.

A central question was whether the neuroendocrine profile in subjects who were not depressed or anxious at the time of the TSST can nevertheless offer indications of their susceptibility to these disorders at other times. Our findings show that this is indeed the case.

In the overall group of subjects who develop depression at another point in the year, the CORT response reached its peak well after the end of the stressor and continued longer than in subjects who did not develop depression, suggesting that stress hyperresponsiveness is indicative of vulnerability. Interestingly, this pattern was especially evident in subjects that developed moderate levels of depression, where the entire area under the curve was significantly increased during the stress response. In the small number of subjects with more severe follow-up depression, we have evidence of a highly dysregulated neuroendocrine profile in response to the TSST, even though these subjects did not meet depression criteria at the time of the test. Thus, while neither groups exhibit “stress fitness”, different endocrine profiles appear to be predictive of different levels of depression severity. Clearly, larger N’s are needed to confirm and extend these observations.

One of the most notable findings in this study is our ability to predict the incidence of anxiety specifically in females. Female subjects who were not currently anxious but who developed anxiety at other time points exhibited elevated CORT levels before, during and after the TSST, suggesting that CORT hypersecretion is indicative of a lack of stress fitness in this group. Moreover, this increase in CORT did not appear to be driven by an increase in ACTH, suggesting the possibility of adrenal hyperresponsiveness, coupled with efficient negative feedback. This is in contrast to males who developed anxiety, where there was a non-significant trend towards higher pre-TSST CORT levels. This suggests that an elevated anticipatory response may be indicative of vulnerability, but more work is needed to confirm this finding.

Importantly, there was no correlation between the neuroendocrine profile and the Affect Score, and this suggests that these are two different dimensions towards predicting depression or anxiety. In the case of females, the risk due to high CORT and high Affect Score was additive in predicting anxiety. Indeed, ∼52% of the females with the combination of high Affect Score and high CORT developed anxiety, compared to ∼5% of females who had low Affect Score and low CORT levels. Females with high Affect Score but normal CORT showed intermediate levels of risk, with ∼30% developing anxiety. In males, we saw a similar, but not as strong of an effect in the case of depression. Therefore, before clinical evidence of anxiety or depression, we observed biological differences in stress responsiveness (CORT response) and psychological differences at baseline (Affect Score). These differences are sex dependent.

Taken together, the study has added to our ability to predict the emergence of symptoms of depression and/or anxiety in a young, healthy population using a combination of psychological and biological measures. While the Affect Score is clearly our strongest and most replicable predictor, we have shown for the first time that the neuroendocrine profile can be predictive in its own right, especially in females who are susceptible to anxiety. Importantly, these two predictors are not correlated, and their combination proved to be highly informative.

Much remains to be understood, including the signature of milder versus more severe forms of depression, as suggested by our preliminary findings on this dimension, and the potential for other markers of lack of stress fitness in males. Importantly, we do not know whether a dysregulated stress response is simply a marker of susceptibility to mood and anxiety disorders, or whether it is a contributing factor in its own right that needs to be targeted in treatment and prevention efforts. Future work will determine whether we can modify the neuroendocrine response by psychological training, such as meditation or cognitive behavioral therapy. The combination of our findings clearly shows that it is possible to identify risk factors, both psychological and physiological in a young people, which opens the door for efforts at preventing the emergence or chronicity of depression and anxiety in this population.

## Data Availability

All data produced in the present study are available upon reasonable request to the authors

## Acknowledgments

This study was supported by the Office of Naval Research Grants N00014-09-1-0598, N00014-12- 1-0366 and N00014-19-1-2149, the Hope for Depression Research Foundation, and the Pritzker Neuropsychiatric Disorders Research Consortium Fund LLC (https://pritzkerneuropsych.org). We would like to thank the Michigan Institute for Clinical and Health Research for help with this study (UL1TR002240). A shared intellectual property agreement exists between this philanthropic fund and the University of Michigan, Stanford University, the Weill Medical College of Cornell University, the University of California at Irvine, and the HudsonAlpha Institute for Biotechnology to encourage the development of appropriate findings for research and clinical applications.

