## Supplementary tables and figures for "The Neuroendocrine Profile During the Trier Social Stress Test in College Freshmen Offers Insights into the Emergence of Anxiety and Depression Symptoms"

### Supplementary Material

**Supplemental Table 1:** Demographics for freshmen at the University of Michigan across all years for the TSST subjects.

| | | Percent/Range |
|---|---|---|
| Age | | 18-21 |
| Sex | | |
| | Male | 45.40% |
| | Female | 54.60% |
| Race | | |
| | American Indian/Alaska Native | 0.89% |
| | Black or African American | 2.08% |
| | Asian | 19.88% |
| | White | 70.62% |
| | Native Hawaiian or Other Pacific Islander | 0.89% |
| | More than one race | 3.56% |
| | Other | 2.08% |
| Ethnicity | | |
| | Hispanic | 7.71% |
| Depressed at Baseline | | 4.45% |
| Depressed at Follow-up | | 23.66% |
| Anxious at Baseline | | 6.23% |
| Anxious at Follow-up | | 21.77% |
| Either Baseline | | 9.20% |
| Either Follow-up | | 30.91% |
| Both Baseline | | 1.48% |
| Both Follow-up | | 14.51% |
| Neither at Baseline | | 90.80% |
| Neither at Follow-up | | 69.08% |
| Baseline Ns | | 337 |
| Follow-up Ns | | 317 |

**Supplemental Table 2:** Repeated measures ANOVA for difference in CORT response between all subjects who developed follow-up anxiety.

| | Sum Sq | Mean Sq | Num DF | Den DF | F val | P val |
|---|---|---|---|---|---|---|
| TSST timepoint | 4,548.97 | 379.08 | 12 | 3,197.05 | 43.50 | 0.00 |
| Follow-up Anxiety | 41.95 | 41.95 | 1 | 266.98 | 4.81 | 0.03 |
| Interaction term | 50.57 | 4.21 | 12 | 3,197.05 | 0.48 | 0.93 |

**Supplemental Figure 1:** PHQ-9 at the time of the TSST was correlated with the rise in CORT.

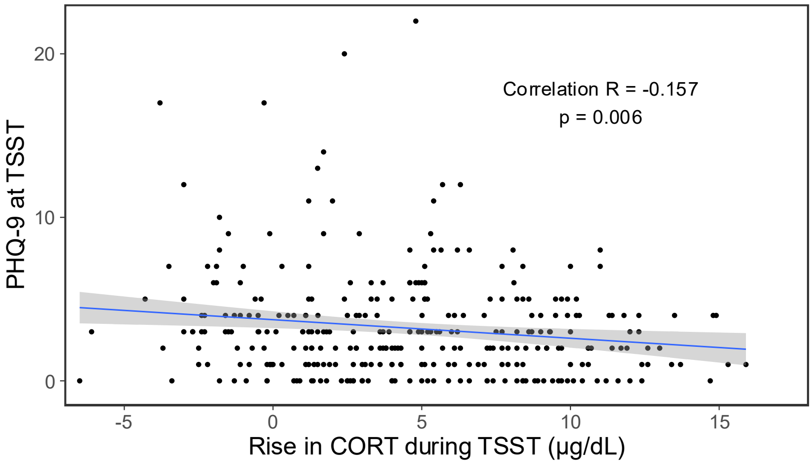

**Supplemental Figure 2:** There were differences in the ACTH (A) and CORT (B) response to the TSST for Control subjects with no depression (PHQ-9 < 10), subjects with moderate depression (PHQ-9 ≥ 10 and PHQ-9 < 15) and subjects with severe depression (PHQ-9 ≥ 15). For Controls, N = 222 (110 female), for moderate depression, N = 36 (22 female), and for severe depression, N = 11 (5 female).

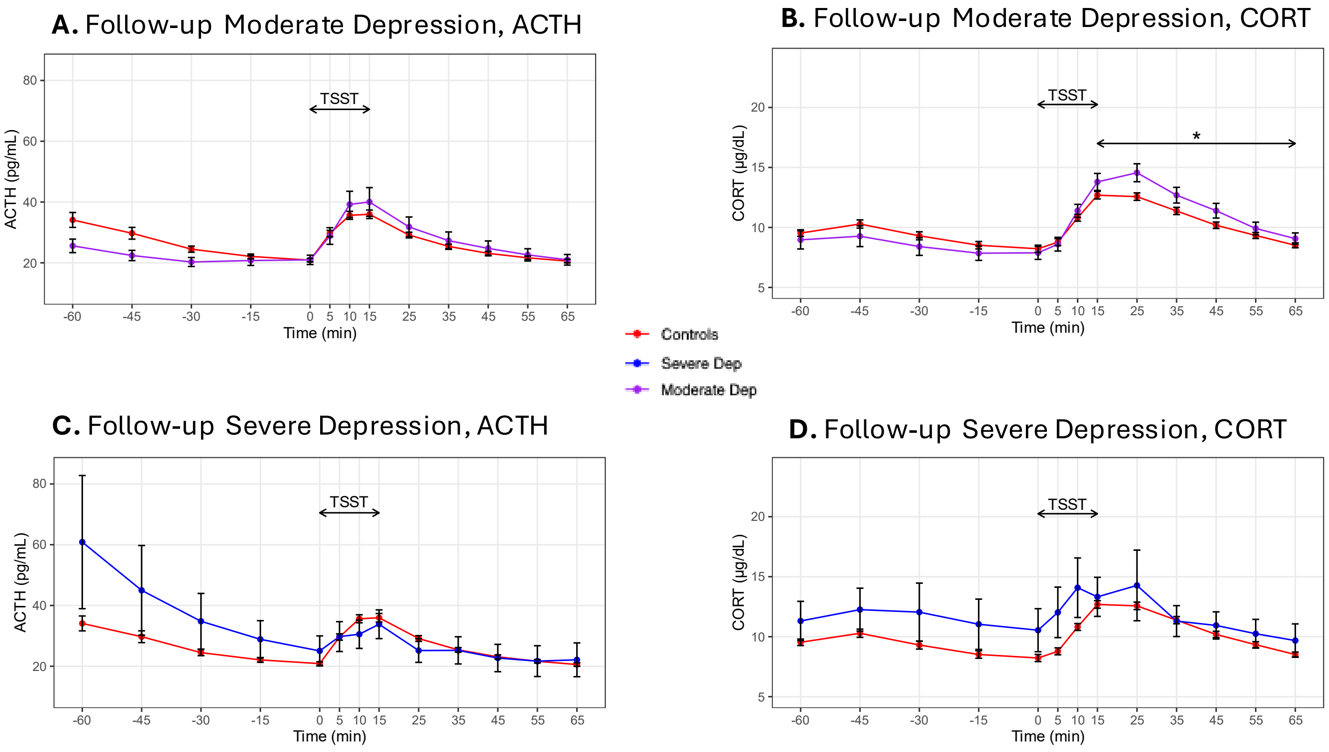

**Supplemental Figure 3:** The effect of high CORT and high Affect is additive in males but not females for depression.

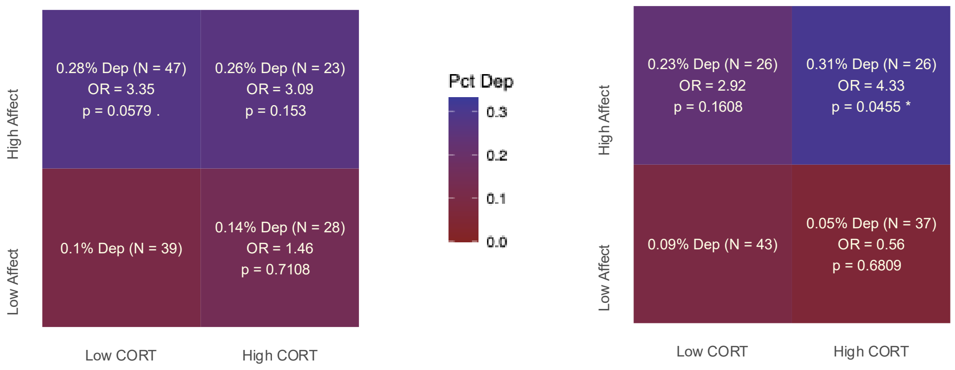
